# Rising to the Ultrasensitive Rapid Diagnostic Challenge with Buoyant-Analyte-Magnetic (BAM) Assays

**DOI:** 10.1101/2024.12.23.24319448

**Authors:** Chuanlei Wang, Emory Satterfield, Carolina B. Livi, Delphine Dean, Jeffrey N. Anker

**Author notes:** Corresponding author email: Jeffrey Anker.

## Abstract

Ultrasensitive, inexpensive, rapid, and portable assays are needed to detect infectious diseases at early stages to improve treatment and prevent transmission, especially to vulnerable patients. Unfortunately, most ultrasensitive assays require rinsing and development steps which increase their complexity, cost, and read time. We address these issues in a novel assay that uses a combination of buoyant microbubbles and magnetic microspheres to doubly label an analyte (SARS-CoV-2 N-protein from surfactant-lysed viruses) and form buoyant-analyte-magnetic (BAM) sandwich complexes. A permanent magnet pulls down the BAM complexes, while unbound microbubbles float towards the surface and separate without rinsing. Removing the magnet releases the buoyant BAM complexes allowing them to float upwards. Under flashlight illumination, they appear as bright rising dots. A simple digital camera (or even cell phone camera) counts the BAM complexes. Remarkably, the analytical detection limit is ∼50 N-protein molecules in 5 µL of saliva. The assay gave positive results for all PCR-positive saliva specimens tested, with concentrations ranging from 0.7 to 2.51×10^5^ RNA copies/µL. We also discuss assay limitations and ways to address them in the future.

## Introduction

At early stages of Covid-19, and many other diseases, specific biomarkers are found in bodily fluids at very low concentrations^1–4^. Detecting these biomarkers early is important both to improve treatment outcomes and prevent transmission, especially in outbreaks and pandemics, which can have devastating effects on public health and the economy. For example, the World Health Organization (WHO) estimated there were 14.9 million excess deaths during the first two years of the Covid-19 pandemic,^5^ and at times many businesses were closed to limit disease spread.

When time is limited and resources or funding are constrained, diagnostic tests should ideally adhere to the WHO’s ASSURED criteria (Affordable, Sensitive, Specific, User-friendly, Rapid/Robust, Equipment-free or battery-powered, and Deployable).^6,7^ Unfortunately, current assays for Covid-19 and many other diseases are either insufficiently sensitive or are insufficiently rapid and affordable (Figure 1). Generally, qPCR meets the sensitive and specific criteria well, but for many applications is not sufficiently affordable, user-friendly, rapid, equipment-free, or deployable^8^. In contrast, the rapid lateral flow assays widely available in pharmacies^9,10^ typically meet six of the criteria (affordable, user-friendly, specific, rapid/robust, equipment-free, and deployable), but their sensitivity is low, with real-world false negatives occurring frequently (∼50% sensitivity for asymptomatic patients and 70% for symptomatic COVID-19 patients)^11^. As a result, these tests often fail to detect infectious diseases at levels that can nonetheless be infectious, especially with prolonged close contact^12,13^. Other portable detection technologies have also been developed for Covid-19, including portable isothermal nucleic acid amplification tests, such as Abbott’s ID Now test. Unfortunately, these devices are somewhat expensive (due in part to costs of equipment, enzyme reagents, and profit), and they have a reported detection limit of 31×10⁵ RNA copies/mL, which means that patients can still be infectious below this threshold^13^. Thus, more sensitive and faster assays are needed for cases requiring a definitive diagnosis (e.g., urgent care facilities)^14^, to prevent transmission to vulnerable patients (e.g., skilled nursing facilities)^15^ and to more effectively contain disease outbreaks^16,17^.

**Figure 1.**
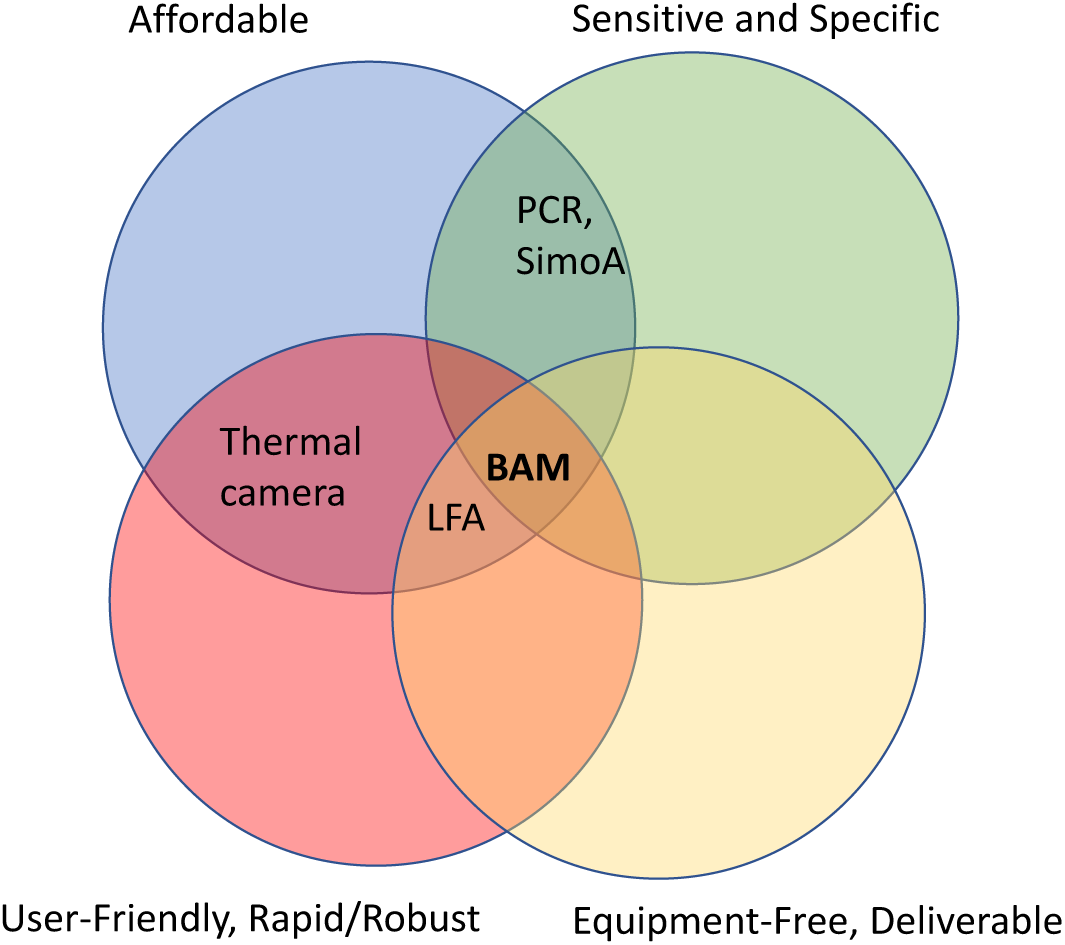
Venn Diagram of the WHO’s ASSURED criteria (Affordable, Sensitive, Specific, User-friendly, Rapid/robust, Equipment-free/battery powered), Deliverable) for BAM assays and other techniques.

The most sensitive antigen tests available label and count single molecules^18^. Their high sensitivity requires both an intense signal (from bright labels that efficiently bind to the analyte) and a low noise level (from instrumental noise and variation in background from unattached or nonspecifically bound labels). To attain this ultra-sensitivity, antigen assays often sandwich the analyte between two types of antibody-functionalized labels: magnetic microspheres and a detection label such as the enzyme horseradish peroxidase (HRP). Using concentrated magnetic microspheres reduces the analyte to capture surface diffusion-reaction time compared to flat surfaces^19^ enabling high capture efficiency in a reasonably short time. The magnetic microspheres are then collected with a magnet and rinsed to remove unbound detection label.

Any detection label still bound to the magnetic microspheres is then enzymatically developed to provide a bright signal^20,21^. For example, Walt’s group developed a single molecule array (Simoa) assay, subsequently commercialized by Quanterix, using magnetic microspheres and HRP enzymes as a fluorogenic secondary label^22,23^. Similarly, Mirkin’s group developed bio-barcode assays using magnetic microspheres and DNA-functionalized gold nanoparticles as the label, with PCR/gene chip or silver development as a readout^24^. However, the required rinsing and development steps add time, protocol complexity, and instrument complexity, which makes it difficult to deploy rapid assays at the point-of-care (POC) or within resource-constrained environments.

Herein, we describe a single-molecule counting magnetic sandwich immunoassay for SARS-CoV-2 nucleocapsid protein (N-protein), which avoids the need for rinsing and development steps, simplifying instrumentation and protocols. Uniquely, we use ∼15 µm gas-filled silica-shell buoyant microbubbles as detection labels in our sandwich assay (Figure 2). These microbubbles have four key advantages: First, the buoyant microbubbles rapidly sample a large solution volume as they rise in solution (∼2 mm/min), enabling rapid and efficient analyte and magnetic particle capture^25^. Thus, buoyant microbubbles have previously been used for high capture efficiency, simple cell separation,^26^ and the separation and detection of *Escherichia coli*^27^, as well as ultrasensitive electrochemical depletion assays.^28^ Second, the unbound microbubbles rise away from the bottom surface during magnetic separation, which clears the background solution in minutes without rinsing. Third, microbubbles and BAM complexes scatter light from a flashlight so intensely that they can be observed by the eye and can be counted using inexpensive portable cameras. Finally, opposing buoyant and magnetic forces reduce nonspecific binding by pulling apart weakly bound BAM complexes and keeping them separated. We employed these features to develop a rapid single-molecule-counting assay using inexpensive portable equipment.

**Figure 2.**
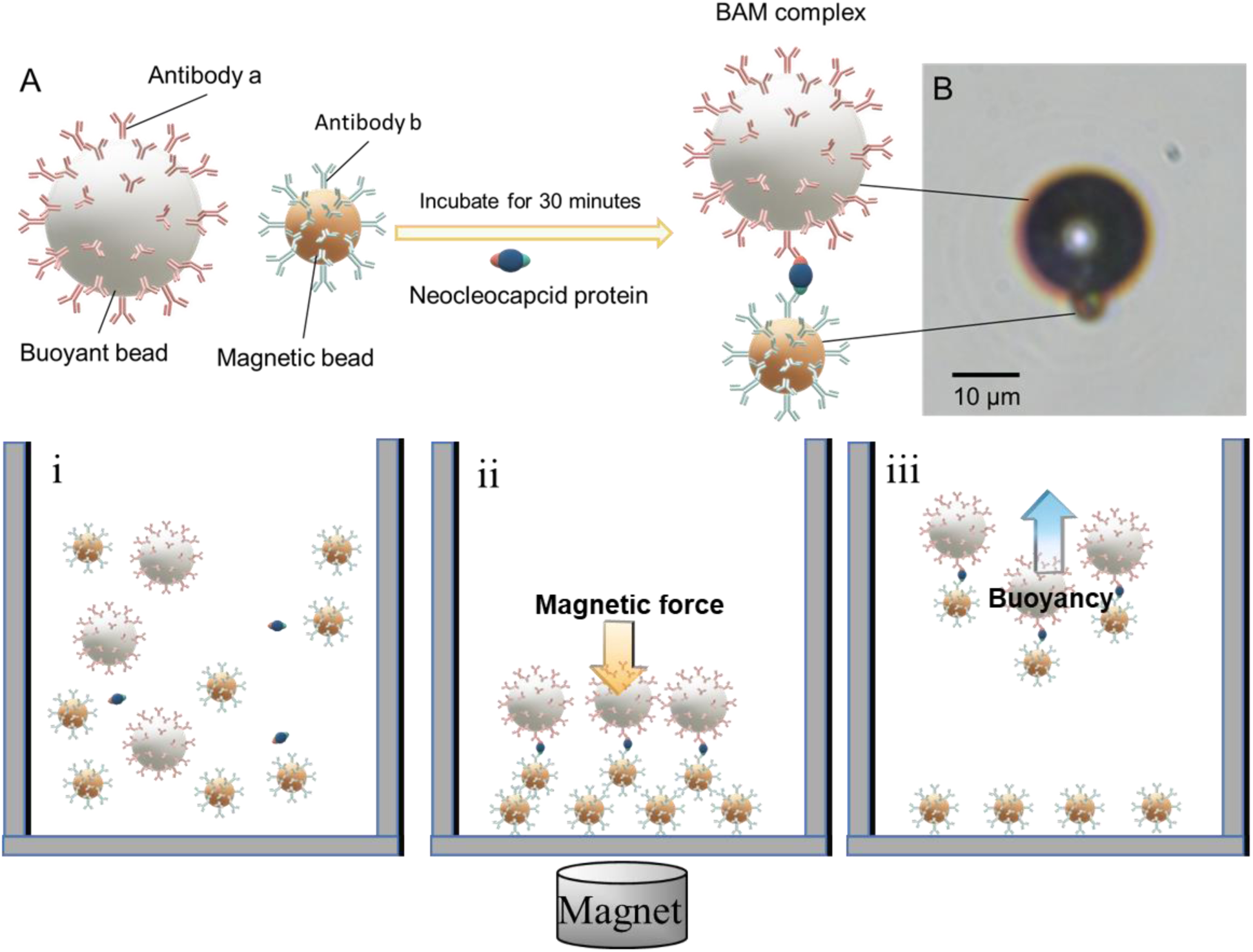
A) Schematic for BAM complex formation. **B)** BAM complex bright field microscopy image. **C)** Illustration of BAM procedure steps (i) Incubation to form BAM complexes. (ii) Magnetic collection. (iii) BAM complex release and counting.

## Results

We first calibrated the BAM assay using simulated saliva samples. We then performed qualitative and quantitative analysis in patient saliva samples and compared results to saliva PCR in aliquots taken from the same specimen.

### Simulated Saliva Studies

Simulated saliva was spiked with varying N-protein concentrations and the BAM complexes were counted. The nonspecific background on simulated saliva with no N-protein was 12.5 ± 8.8 BAM complexes. Figure 3 illustrates that the linear regression model provides an excellent fit to the data, with an R-squared value of 0.99. The slope of 0.84 BAM complexes per molecule suggests an analyte capture-and-detection efficiency of 84%. The analytical limit of detection (LOD) was 31 N-proteins, given by 3s_y_/m, where s_y_ is the noise on the blank (8.8 BAM complexes) and m is the calibration curve slope, (0.84 BAM/N-protein). This is equivalent to 0.24 fg/mL in 10 μL 10% simulated saliva, or 2.4 fg/mL in the original 1 µL saliva before dilution to 10%.

**Figure 3:**
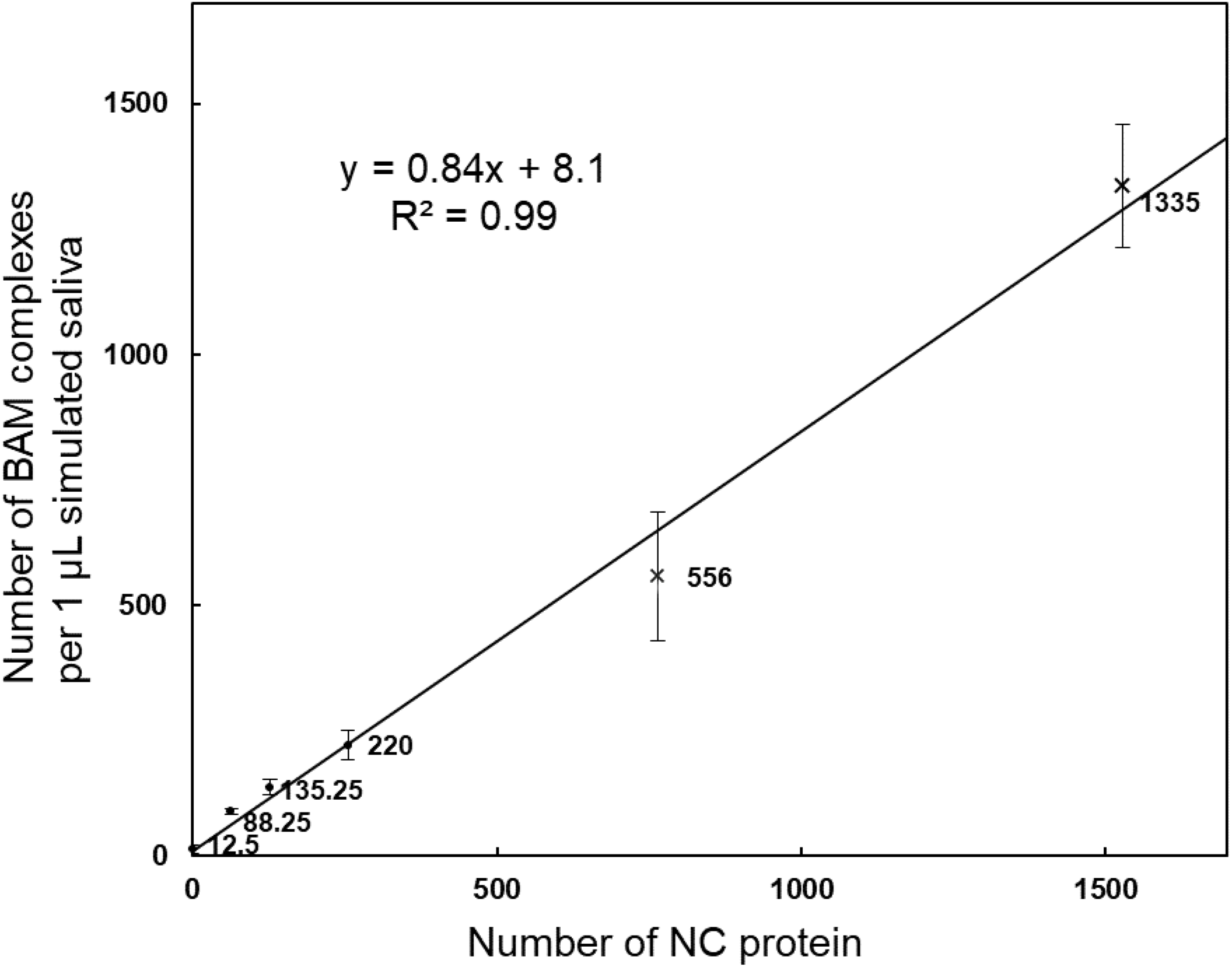
Calibration curve for SARS-CoV-2 N-protein molecules in 10% simulated saliva. An average of 0, 64, 127, 255, 764 and 1528 N-protein molecules were spiked into simulated saliva and resulting BAM complexes were counted. At above 255 molecules/μL, BAM complexes were hard to count directly due to overlapping tracks; instead, BAM complexes were counted in 1/10 of the mixture and multiplied by 10x. Each point shows the average and standard deviation of 4 trials.

In addition to quantitative results, several qualitative observations were made. Within 15 minutes of applying the magnetic field, all magnetic particles were at the bottom of the cuvette, and most of the buoyant microbubbles had clarified off the bottom, except for some occasional low-buoyancy debris. Additionally, although not part of the standard protocol, we found that BAM complexes remained intact after releasing the magnetic field and could be retracted back to the cuvette bottom by reapplying the magnet. Furthermore, we found that during the second pulldown, the background further decreased as particles spontaneously pulled apart. The reduction decreased after each pulldown (e.g., see Supplementary information, Figure S1). We also determined that surface chemistry is critical in maintaining low background levels, as a significantly larger background was observed without biotin-PEG functionalization (Supplementary information Fig S2A). Lastly, after performing BAM assays after various incubation times, the exponential regression line fit well with a 8-minute time constant (Supplementary information, Figure S3 and Table S1). Despite this, a 30-minute incubation was used to ensure high capture efficiency and negligible sensitivity to reaction rate.

### Real saliva assays

In our negative real saliva BAM assay, we observed a nonspecific background of 77 ±10 BAM complexes (Supplementary information Figure S4). This background, although relatively low, was significantly larger than the background in simulated saliva, which is likely due to mucin and other molecules found in saliva. Diluting the saliva by 2x, 10x, and 100x, respectively, reduced the background to 36 ± 3, 29 ± 2, and 19 ± 4 BAM counts (Supplementary information Figure S4). Capture-and-detection efficiency was 78% in saliva samples diluted to 1% concentration (Supplementary information, Figure S5D and 91% in samples diluted to 50% concentration (Supplementary information, Figure S6D). Since diluting patient samples to 50% (i.e., 5 µL of pure saliva diluted to 10 µL) provided a good balance between minimizing the negative background and maintaining a high capture efficiency, it was decided that all subsequent experiments would employ a 2-fold dilution.

### Positive and Negative Patient studies

Next, we performed BAM assays on n*=*3 PCR-positive and n*=*8 PCR-negative patient saliva specimens. As shown in Figure 4A, the positive specimen is clearly distinguishable from the negative specimen by the significant difference in the number of BAM tracks. Across all tested specimens (Figure 4B), the average number of BAM complexes in positive samples was 411 ± 151, compared to 19 ± 10 in negative samples. The positive and negative specimens were completely distinguished (p=5×10^-5^) with no negative specimens reading over 49 BAM counts and no positive specimens reading below 208.

**Figure 4:**
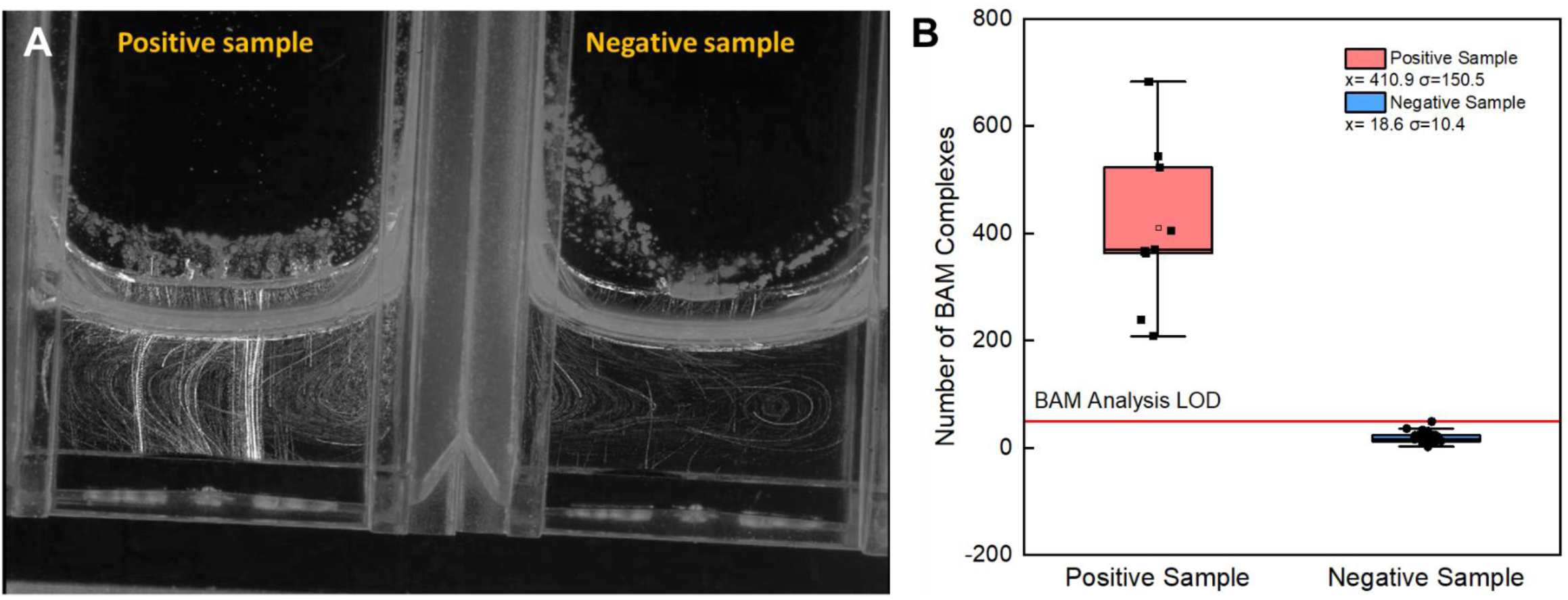
A) BAM track for PCR-positive patient saliva sample in the left cuvette with 216 tracks; negative saliva sample in the right cuvette with 12 tracks. B) Box plot for PCR-positive and negative specimens.

### Serial Dilution and Quantification

The results indicated no increase in the number of BAM complexes following the addition of concentrated N-protein solution, suggesting that the hook effect, which states that very high levels of analyte can lead to a paradoxical decrease in assay signal, was not observed. This result illustrates that the assay loses linearity at high N-protein concentrations (see supplementary information Figure S7A).

To address the difficulties counting >∼250 BAM complexes due to overlapping tracks, and non-linearities from aggregation, and surface saturation, we diluted each specimen until it was in a linear range. Further diluting the diluted PCR-positive samples by an additional 100x (up to 10^8^-fold overall), resulted in 20 ± 9 BAM complexes, closely matching the negative background from PCR-negative samples (19 ± 10 BAM complexes, Figure 4B).

After accounting for the dilution factor (for the solutions diluted to the linear range), the BAM concentrations were compared to RNA concentrations calculated from C_t_ values (Figure 5). The slope of the best-fit line suggests approximately 470 BAM complexes per RNA copy (Figure 5A). This linear relationship between BAM complexes and RNA is maintained over a wide concentration range, from <1 cp/µL to 25000 cp/µL, levels high enough to be detectable by LFA (Figure 5B).

**Figure 5:**
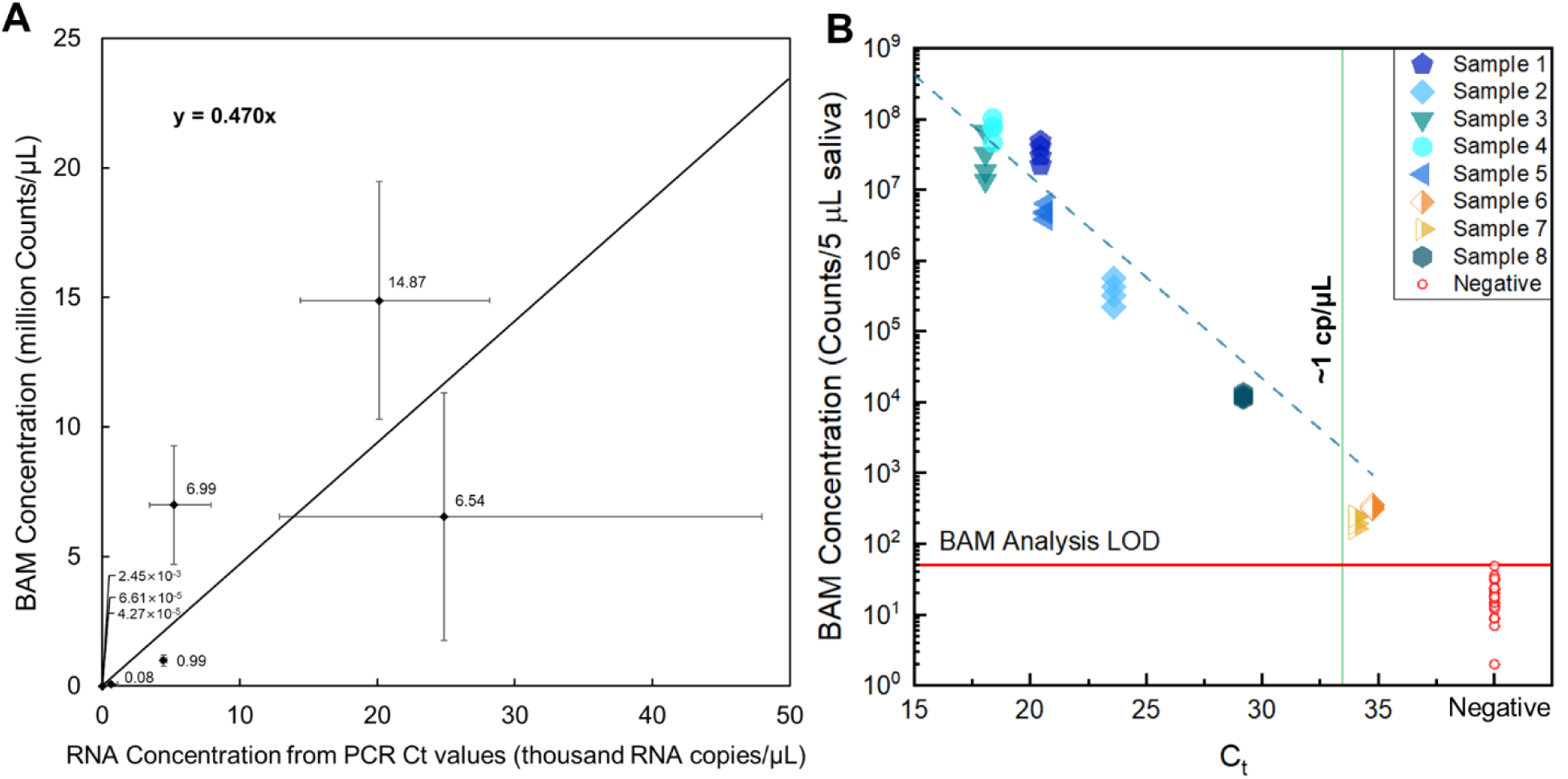
Comparison between BAM counts (after lysis and serial dilution) vs. PCR-measured RNA copies/μl for n=8 PCR positive and n=8 PCR negative patient saliva specimens. Although all positive specimens gave positive BAM results at initial concentration, overlapping tracks prevented quantification; thus, for quantification, specimens were diluted by up to 106x to the linear assay-response region. **A)** Plot on linear scale. **B)** Plot from A on a log yscale and PCR Ct value x-scale. The linear regression slope was ∼470 BAM complexes/RNA. The analytical LOD from negative specimens was 50 BAM complexes in 5 μL patient saliva.

## Discussion

The BAM assay was designed for ultrasensitive rapid affordable detection of early-stage diseases. Below we discuss the assay’s LOD, quantification, protocol time, affordability of equipment and disposables, and deployability/user-friendliness.

### LOD

The analytical signal LOD for the patient saliva assay was 37 N-protein molecules per 5 µL, or 0.29 fg/mL, or 12 aM. To the best of our knowledge, this is the lowest LOD for N-protein concentration reported to date. For example, Quanterix commercialized a single molecule array (Simoa) assay with antibody-functionalized magnetic microspheres to capture the N-protein and an antibody-functionalized detection enzyme to develop it. The LOD in nasopharyngeal swabs diluted in 100 µL buffer was reported as 99 fg/mL,^29^ and an in-house Simoa assay was reported with an LOD of 20 fg/mL^30^. Other sandwich immunoassays with magnetic separation for other proteins have LODs almost as good. For example, Jwa-Min Nam et al., report a 30 aM LOD for human prostate-specific antigen (PSA) in 10 µL of goat serum using a biobarcode sandwich assay with a DNA-detection label and PCR as a readout^24^. However, the BAM assay does not require rinsing and development steps, which simplifies protocols, enables use of less expensive, more portable instruments, and can reduce minimum total assay time.

The BAM assay’s ultralow LOD is due to both high capture-and-detection efficiency (in 30 minutes, 78-95% capture-and-detection efficiency) and low noise from variation in blank samples. Each BAM complex scatters light intensely, enabling high detection efficiency. The lack of rinsing, nanowell loading, or development steps also avoids lost. The high analyte capture efficiency could be explained by the large volume explored by the microbubbles as they rise. For example, Literature on planktonic feeding shows that rising at ∼50 µm/s increase the rate of analyte molecule capture ∼2x compared to diffusion,^31^ and greatly increases the rate of subsequent capture of magnetic particles (which diffuse slowly due to their size).^32,33^

In addition to capturing and detecting most molecules, the excellent LOD arises from low nonspecific binding background variation. The background from n=8 negative patients (24 test) was 19 ± 10 (Figure 5B), and the background from the same number of positive specimens, after diluting by up to 10^6^-fold was 20 ± 9 (Supplementary information Table S2). The total background counts are low, and variation is correspondingly low, although it was higher than shot noise and included variation from multiple specimens, days, and lots. Moreover, the assay consistently detects very low number of molecules, including in calibration curves (Figs S5D and S6D), the reaction rate curve (Supplementary information Figure S3 and Table S1), and correctly identified 8 negative specimens, 32 positive trials, another n=8 positive specimens during serial dilution by up to a factor of 10^6^. More studies are planned after we develop the next generation portable prototypes and rapid protocols.

Mechanistically, two factors contribute to the low nonspecific binding background. First, good surface chemistry and blockers are essential (e.g., Supplementary information Figure S2) shows that without biotin-PEG to block free streptavidin sites on the buoyant and magnetic microspheres without antibodies, significantly more background is observed). Second, the opposing buoyant and magnetic forces can break apart weakly nonspecific interactions and keep them separated without significantly decreasing strong interactions including specific bonds. We estimate a 15 µm microbubble with a density of 0.6 g/cm^2^ has a buoyancy force in water or buffer of ∼7 pN. Since the BAM complexes are pulled down by the magnet around 5x as quickly as they rise, we expect that during this process the magnetic and buoyant forces are pulling on the N-protein with ∼6x as much force or ∼40 pN, albeit for the brief period (∼30 s) when the BAM complexes close to the bottom of the cuvette but haven’t yet reached it). Previous studies on pulling apart bonds with AFM tips, magnetic tweezers, centrifugation, and optical tweezers show that the time to dissociate decreases exponentially with time, roughly in accordance with Bell’s model.^2–4^ The forces and times calculated here are sufficient to keep apart spontaneously-dissociating particles and break apart weak interactions, but over the short experiment, are unlikely to affect antibody-antigen interactions with high K_D_ (e.g., 10 pM for the R040 anti-nucleocapsid antibody) and force-free dissociation times of days to months.^34,35^ The forces were sufficient to reduce nonspecific binding while retaining high capture efficiency; in future the magnetic forces, and bead sizes and surface chemistry can be further optimized.

### Quantification

BAM concentrations were quantified by diluting the saliva specimen to the linear region (and subtracting the 20 BAM complex average background signal and divided by the dilution factor). This quantification approach is similar to how bacteria are traditionally quantified by diluting to a point where individual bacterial colony forming units can be counted for bacteria (or similarly, plaque forming units for viruses). The slope of Fig 4B was 470 BAM complexes per RNA copy, or around 500 N-proteins/RNA, assuming an 84% capture efficiency. This is roughly consistent with electron microscopy of individual (pleomorphic) virions giving 38±10 ribonucleic acid complexes per virion, with simulations of showing 12 N-proteins per ribonucleic acid complex which gives a total of 456±120 N-proteins/virion^36,37^. However, the BAM assay does not directly count N-proteins per RNA, and differences can arise if saliva RNA or N-proteins degrade, if there are free saliva N-proteins not in a virus, or saliva antibodies mask the N-proteins, and due to other noise sources. Indeed, other studies found >10x specimen to specimen variation at the same C_t_ value^29,30^. Overall, the dilution approach provided a robust quantitative analysis over a wide range of concentrations (up to 10^8^ BAM/5 µL saliva).

Admittedly, the serial dilution steps needed for quantification make the assay less rapid, user-friendly, and affordable. If needed, we expect that dilution can be automated with microfluid systems, with different regions in one or more cuvettes used to assess different concentrations.^38^ Additionally, for faster assays (when capture efficiency becomes strongly time-dependent), ratiometric measurements could be performed by multiplexing based on microbubble size, shape, fluorescence, or chemically or photocleavage release time.^39^ However, for most infectious disease applications, quantification is unnecessary and qualitative infection status results would be sufficient, especially for rapid on-site tests.

### Total Assay Time

The current assay protocol was designed to show proof-of-principle, and we used relatively long incubation time (30 minutes) to capture most analyte in the specimen both to improve reproducibility and show excellent LOD. For many point-of-care applications, 30 minutes would be too long, and the need for rotating the sample during incubation would be impractical. However, we found that the BAM formation time constant was 8 minutes (Supplementary information Figure S3 and Table S1). Thus, theoretically a 1-minute incubation time should provide 10% capture efficiency, allowing manual mixing (e.g., by squeezing a tube), which would be sufficient for many applications.

The total assay time also includes time for saliva collection, magnetic separation, and readout with the magnet removed. The collection step was ∼ 4 minutes including Salivette centrifugation but could be speeded especially for small 5 µL specimens. We used 15 minutes for magnetic separation, and this was needed to have a clear region into which the BAM complexes rise, but if the small, slowly rising microbubbles were removed prior to running the assay, this time could be reduced. Finally, we recorded up to 10 minutes of readout after the magnet was removed to be able to analyze BAM particle and background motion, but over half of the BAM complexes could be counted within 10 frames (20 s), (Supplementary information Figure S9 and Table S3), and larger beads would be detected even sooner. Thus, although the total assay time was around 55 minutes, we expect that it could be reduced to ∼5-15 minutes, with some loss in LOD but still unprecedented sensitivity for a rapid portable immunoassay.

### Affordability of instrument and disposables

Importantly for meeting the WHO’s ASSURED criteria, the equipment and off-the-shelf small components and reagents were all affordable/inexpensive. The most expensive and heaviest component is the Nikon D7000 DLS camera. However, since the particles are visible by eye, most digital cameras will work, and indeed we found a phone camera with no additional optics (Galaxy flip) could easily detect 1 fg/mL (see Supporting Information Figure S10 and movie M2). In future, we are exploring this and other alternatives, such as coin microscopes, which are widely available for $20-60. The remaining equipment components (flashlight, magnet and cuvette holder) is relatively small and inexpensive and expect these could be manufactured for <$5.

Regarding disposable costs, very little reagent is used per test. The main costs are for the Sailivette saliva collection swab ($0.50/test), cuvette ($0.10/test), buoyant beads + antibody functionalization $0.32/test, magnetic beads + antibody functionalization $0.15/test. This gives a cost-per-test of $1.07 at list prices. However, there are some minimum order costs, and we estimate that $6,200 is enough to buy reagents and cuvettes for around 3,600 tests, $1.72 per test (Supplementary Information Table S4). In practice, including validation standards, and costs for storage/expiration, shipping, packaging, labor, and business expenses would raise total costs, while mass production and protocol cost optimization would lower them. Regardless of the precise cost, BAM tests can potentially meet affordability and deliverability in the ASSURED criteria.

### Deployability and User-friendliness

This paper presents proof-of-principle for ultrasensitive BAM assays. To move towards real applications, we will need to make the instrument smaller and lighter (e.g., using a coin camera or cell phone). We will also have to simplify and automate the protocol to collect saliva, mix reagents, read results and avoid accidental spillage. Reagent storage and shelf-life will require proper selection of antibodies or antibody fragments, covalent attachment to the bead, and lyophilization (possibly in a cassette or pad) to enhance antibody stability. Additionally, regulatory approval on a finalized product would be required for clinical use.

## Conclusions

This article describes the first single-particle BAM assay and applies it to the simple, fast, cost-effective, and ultrasensitive detection of SARS-CoV-2 N-protein in saliva. This test bridges the gap between PCR and rapid antigen tests by significantly enhancing the sensitivity and specificity of antigen tests while maintaining their ease of deployment. Unlike traditional rapid tests, which often sacrifice accuracy for speed and simplicity, this method combines the high sensitivity typical of PCR with the rapid, low-cost nature of antigen testing. By leveraging buoyant microbubbles for precise analyte detection, the BAM assay offers a portable solution that doesn’t require complex lab equipment or extensive training, making it highly suitable for point-of-care diagnostics in a variety of settings.

What sets this method apart is its use of approximately 15-µm silica-shell microbubbles as labels. These microbubbles are buoyant, allowing for the rapid and efficient formation of BAM complexes with extremely low backgrounds. The buoyant properties of the beads also automatically separate unbound labels from BAM complexes, which are large enough to be observed with the naked eye and counted using inexpensive portable cameras. BAM assays can utilize affordable and portable equipment, although further optimization is required to enhance ease of use and speed.

Future work involves simplifying the sample collection and optimizing the protocol for user-friendliness, including developing integrated portable setups, automating the particle counting and analysis, testing in larger patient trials, and extending the approach to other disease-specific analytes.

## Online Methods

Our method utilizes antibody-functionalized buoyant microbubbles and magnetic microspheres to capture and detect single molecules (Figure 2A). This approach is based on research by McNaughton et al., employing BAM for the capture and separation of *E. coli*^40^. Our objective is to perform quantitative experiments on the analyte during the BAM complex separation stage by ensuring that the BAM complexes possess both magnetic and buoyant properties. In the initial step of this immunological analysis method, magnetic microspheres and buoyant microbubbles form sandwich antibody complexes. Assuming capture is stochastic and beads are roughly uniform in size, at low concentrations, the number of molecules captured per microbubble follows a Poisson distribution.^23^ When a large number of microbeads are used to capture analytes at extremely low concentrations, only a small fraction of the microbeads are labeled, with the vast majority remaining unlabeled.

### Materials

The magnet used in the BAM assay was made from a neodymium iron boride alloy with two shapes. The magnet disc (cat. D1051A) has a diameter of 8 mm, a thickness of 4 mm, and a Neodymium strength of 45. The magnet cylinder (cat. Cyl0164) has a diameter of 6 mm, a height of 8 mm, and a Neodymium strength of 50. They were both obtained from SuperMagnetMan (Pelham, AL, US). The LED light (Cat rechargeable extendable LED work light, cat: CT3115) was purchased from EZRED (Denville, NJ, US). BSA powder (Bovine Serum Albumin, A7030-10G) was obtained from Sigma-Aldrich (St. Louis, MO, US). The semi-millimeter cuvette (product name: BRAND® semi-micro cuvette, cat: BR759115) was purchased from MilliporeSigma (Burlington, MA, US). Low protein binding microcentrifuge tubes (Cat: 90401), Gibco PBS pH 7.2(Cat: 20012027) and the Pierce Antibody Biotinylation Kit for IP (Cat: 90407) were obtained from Thermo Scientific (Rockford, IL, US). A 66 mm width, 500 mm length optical rial (cat: XT66-500) and two 66 mm “Clamping Platform with Counterbored Slot” (cat: XT66C4) were obtained from Thorlabs (Newton, NJ, US). Streptavidin Microbubbles (buoyant beads) were purchased from Akadeum Life Science (Ann Arbor, MI, US). The magnetic beads were LodeStars Streptavidin 2.7 μm Magnetic Beads, which were provided by Agilent (Santa Clara, CA, US). The SARS-CoV-2 nucleocapsid protein (Cat: NUN-C5227-100ug, a 47.3 kDa protein synthesized with a his-tag) was obtained from Acro biosystems (Newark, DE, US). The biotin-mPEG 5K (Cat: PLS-2054) was obtained from Creative PEGworks (Durham, NC, US). The Triton-X 100 and carboxymethylcellulose sodium salt were obtained from Alfa Aesar (Ward Hill, MA, US). All aqueous solutions used in the experiments were prepared using Milli-Q water (18.2 MΩ/cm) obtained from a ELGA Purelabflex2 system (ELGA LabWater, Woodridge, IL, US). The chemicals for lab-made, artificial saliva samples were as follows: Sodium chloride (NaCl) was obtained from Spectrum Chemicals (New Brunswick, NJ, US). Calcium chloride (CaCl_2_), magnesium chloride (MgCl_2_) and potassium phosphate dibasic (K_2_HPO_4_) were obtained from Sigma-Aldrich (St. Louis, MO, US). The carboxymethylcellulose sodium was obtained from Alfa Aesar (Ward Hill, MA, US). Potassium chloride (KCl) was obtained from Thermo Scientific (Rockford, IL, US). And the urea was obtained from J.T. Baker (Phillipsburg, NJ, US).

#### Experimental setup

As shown in Figure 6, the experimental setup consists of a 3D printed magnet slide holder and cuvette holder, six magnet discs, two magnet cylinder, a digital camera (Nikon D7000), two LED light sources, and a 66 mm optical rial (according to the materials section above). The upper end of the semi-milliliter cuvette was slightly tilted (3°) towards the camera direction and fixed onto the cuvette holder. The small tilt angle prevents BAM complexes from touching or getting stuck on the cuvette walls as they rise. The setup includes a total of six magnetic discs and two magnetic cylinders, with each set comprising three magnetic discs and one magnetic cylinder. At the bottom of the magnet slide holder, two magnetic discs are positioned, while one magnetic disc and one magnetic cylinder are placed on top. When placing or removing the magnets, moving the magnet slide allows for the simultaneous placement and removal of both sets of magnets. Subsequently, the LED lights placed on both sides of the cuvette were turned on, and the camera position was adjusted on the optical rail until a clear image of the two cuvettes appeared in the camera view.

**Figure 6:**
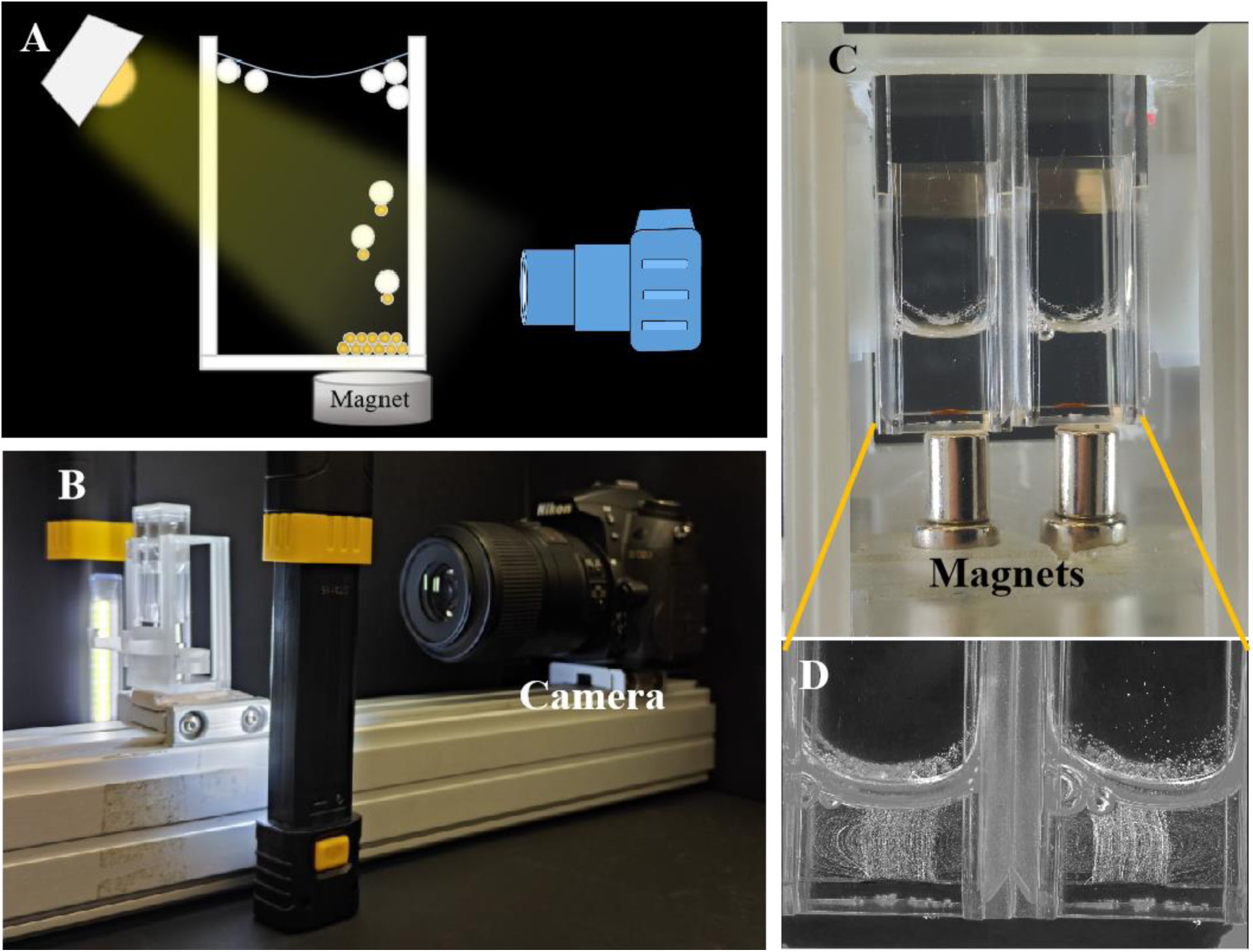
(A) The schematic illustration for BAM test set up including a camera, a cuvette holder fixed on the optical rial. The cuvettes are placed on the cuvette holder with magnets under the cuvette. Two light sources are placed aside from the cuvette. (B) Picture for BAM test set up. (C) Zoom in figure for the video recording zone. (D) Tack analysis gray map result for BAM test.

### Protocols

#### Antibody biotinylation

The antibodies immobilized on the microbubbles were SARS-CoV-2 Nucleocapsid antibody, Rabbit Mab (40143-R004, referred to as antibody (R004) below). The antibodies immobilized on the magnetic microspheres were SARS-CoV-2 nucleocapsid protein antibody, Rabbit Mab (40143-R040, referred to as antibody (R040) below). To biotinylate the nucleocapsid protein antibodies (Ab-R004 and Ab-R040), the “Pierce Antibody Biotinylation Kit for IP” was utilized, which reacts with amine groups on the antibodies through an NHS-PEG_4_-biotin linker^41^. The antibody biotinylation kit includes a microtube containing solid NHS-PEG4-biotin, Zeba Spin desalting columns, and 10 mL of 20 × PBS solution. Firstly, dilute the 20 × PBS solution to 1 × PBS (10mM sodium phosphate, 0.15 M NaCl, pH 7.5) and add 100 μL of 1 × PBS solution to the microtube containing solid NHS-PEG4-biotin to prepare an 8.5 mM solution. Modifying 50 μg IgG antibody was calculated to require 1.6 μL of 8.5 mM NHS-PEG_4_-biotin solution at a 40-fold biotin linker concentration. Then, 50 μL of 1 μg/μL Ab-R004 and Ab-R040 were separately placed into 2 mL low protein-binding microcentrifuge tubes, followed by the addition of 1.6 μL of 8.5 mM NHS-PEG4-biotin solution and 48.4 μL of 1 × PBS solution. The mixture was incubated at room temperature for 30 minutes. Simultaneously, two Zeba Spin Desalting Columns were prepared by removing the bottom closure and loosening the top cap. The Zeba Spin Desalting Columns were placed in separate 2 mL microcentrifuge tubes and centrifuged at 1500 × g for 1 minute, discarding the solution in the centrifuge tubes.

This step was repeated twice by adding 300 μL of 1 × PBS solution, centrifuging at 1500 × g for 1 minute, and discarding the solution. Subsequently, the incubated antibody solutions were added individually onto the compact resin bed of each Desalting column, placed in a new low protein binding microcentrifuge tube, and centrifuged at 1500 × g for 2 minutes. At this point, the antibody concentration in the solution was 0.5 μg/μL. The biotinylated NC antibodies were then diluted to 0.05 μg/mL by 1 × PBS solution and stored at –20 ℃.

#### Microbeads surface modification

100 μL of the streptavidin-coated buoyant microbubbles and 12 μL of the streptavidin-coated magnetic microspheres, were added to separate low protein binding microcentrifuge tubes to functionalize them with biotinylate antibody and biotinylated PEG. 2.6 μL of biotin-antibody (R004) was added to the buoyant microbubble solution, and 2.6 μL of biotin-antibody (R040) was added to the magnetic microsphere solution. The solution was pipetted rapidly up and down to mix. After antibody modification, 100 μL of 0.01 mM biotin-mPEG 5K, was added to each centrifuge tube to bock the empty streptavidin site, and the solution was pipetted rapidly up and down to mix. Then, 300 μL of 1 × PBS was added, and were centrifuged at 400 × g for 3 minutes. The subnatant was aspirated from the buoyant microbubble solution, and the supernatant was aspirated from the magnetic microsphere solution. This washing process was repeated twice. After washing steps, a 1 × PBS solution containing 2% bovine serum albumin (BSA) and 1% Triton-X 100 were added, and incubated overnight at 4 °C. The microbeads were washed thrice with 400 μL, 1 × PBS under centrifugation 400 × g for 3 minutes. Finally, the buoyant microbubbles and magnetic microspheres were stored in 400 μL of a 1 × PBS solution containing 0.5% BSA, 0.1% Triton-X 100, with a concentration of 2.5×10^4^ particles/μL at 4 °C. These were used within 24 hours of functionalization.

The lyophilized SARS-CoV-2 nucleocapsid protein for the spiking saliva sample was reconstituted by 400 μL 1 × PBS with a concentration of 250 μg/mL. It was then diluted to the desired concentration with 1 × PBS solution. The final concentration of each bead was approximately 750,000 beads for each test, meaning 750,000 beads per 30 μL of solution.

#### BAM test protocol

30 μL of each antibody-modified buoyant microbubbles and magnetic microspheres was transferred into a 2 mL centrifuge tube using a micropipette. To this, 10 μL of the N-protein solution was added to the tube, and the mixture was incubated on a rotator at room temperature for 30 minutes. After incubation, 130 µL of 1 × PBS was added to the mixture and transferred from the centrifuge tube to a semi-milliliter cuvette, where the mixture was then placed in the 3D-printed cuvette holder. The magnet was placed under the cuvette to collect the BAM complexes. After 15-minutes, the permanent magnets were slid into place below the cuvette to pull down the magnetic microspheres and BAM complexes, while the unbound microbubbles floated upward towards the surface. The magnet was then removed, and the trajectories of the BAM complexes were monitored through timelapse photography. We connected the Nikon D7000 digital camera to the laptop using a USB cable and controlled the camera through the Nikon Camera Control Pro 2 version 2.35.1 software. The camera settings were configured with a shutter speed of 1/1.3 seconds, an aperture of f/40, and ISO 250. In the Interval Time Shooting mode, we set the total number of shots to 310 with a 2-second interval. The choice of 310 shots allowed us 20 seconds to remove the magnet from the cuvette holder. Subsequently, all the photos were transferred to Microsoft’s Video Editor software., where the 300 photos were transformed into a 10-second video with a frame rate of 30 FPS. A MATLAB script was developed to visualize the trajectories of the BAM complexes through every frame of the video (Supplementary Information Figure S8, S9, S11 and Table S3 for the code and processing results).

#### Human saliva collection protocol

The saliva collection protocols were approved by the Institutional Review Boards (IRB) of Clemson University (IRB2021-0703) and Prisma Health (Pro00100731). All participants provided their written informed consent to provide a saliva sample. Participants provided 1-2 mL of saliva collected in a 50 mL conical tube and some demographic data (age, gender, race/ethnicity) to the research study.^42^ Samples were deidentified striped of personal identifying information before being passed to the research team. Part of the samples (200 μl) was used for a RT-PCR clinical diagnostic test^43,44^ and eventual sequencing for SARS-CoV2 variant identification.^45,46^

#### Patient saliva samples treatment

The centrifuge tubes were thawed and opened within a glove bag (No. 690323) from NPS (Green Bay, WI, US) in a biosafety hood (Class II type A2) from Labconco (Kansas, MO, US). To lyse the viruses in saliva and release the encapsulated N-proteins, 400 μL of saliva sample was mixed with 400 μL of 1% Triton-X 1× PBS solution by pipette in the glove bag. Welch, S. R. Welch and K. A. Davies et al. report 5.9 logs reduction in 2 min with 0.5% Triton-X.^47^ We applied the mixture to a Salivette cotton swab Salivette(REF: 51.1534 from Sarstedt AG (Nümbrecht, Germany)), and expressed the fluid by centrifuging in the Salivette kit centrifuge tube at 1000 × g for 2 minutes to filter and remove any debris. The saliva supernatant was collected for testing. Processed saliva samples that have been stored at freezing temperatures may experience flocculation upon thawing. The samples were centrifuge at 400 × g for 5 minutes to collect the supernatant for BAM testing.

#### Dilution of the patient saliva supernatant

The supernatant was transferred to a low protein binding centrifuge tube, where it was diluted with a 0.5% Triton-X 100, 1× PBS solution. In each subsequent serial dilution step, the solution was diluted tenfold, with the pipette tip replaced with each dilution.

#### Simulated Saliva Studies

The artificial saliva used in these studies was artificially made using the following: 15.6 mM NaCl, 16.5 mM KCl, 1.01 mM CaCl_2_, 0.361 mM MgCl_2_, 2.07 mM K_2_HPO_4_, 16.3 mM urea, and 5.0 g/L carboxymethylcellulose sodium salt in one liter aqueous^48^. 10 μL of 10% simulated saliva (1 × PBS) was spiked with N-protein. Each BAM test was run per the protocol discussed under section, “The BAM Test Protocol.” Final concentrations of N-protein in the 10 μL of 10% simulated saliva varied between 0 and 12 fg/mL, with each 1 fg/mL corresponding to 127 N-proteins. In the low-concentration samples (0-2 fg/mL), relatively few BAM complexes were seen, which could be easily counted. However, in the more concentrated samples (6 and 12 fg/mL), the large number of BAM complexes resulted in overlapping tracks, which made it difficult to count. To combat this, the samples were thoroughly mixed and one-tenth of their total volume was taken as a representative sample to estimate the total number of BAM complexes in the entire sample.

#### Real Saliva Assays

These studies were conducted on spiked negative saliva samples. These samples were collected by thawing the stored frozen specimens and pipetting onto a Salivette swab. To remove debris, the swab was then placed into a centrifuge (provided in the purchased Salivette swab kit), note that squeezing with a syringe also works. Without this step, it was found that the number of nonspecifically bound BAM complexes rose significantly. These steps mimic either future direct Salivette collection or integrated filtration steps. Each BAM test was run per the protocol discussed under section, “The BAM Test Protocol.”

#### Positive and Negative Patient Studies

These studies were conducted on n =3 PCR-positive and n = 8 PCR-negative saliva specimens. Because our setup allows for two samples to be tested simultaneously, a PCR-positive specimen (left cuvette) was run alongside a PCR-negative specimen (right cuvette) (Figure 3A and Supporting Movie M1). Each BAM test was run per the protocol discussed under section, “The BAM Test Protocol.”

#### Serial Dilution and Quantification Studies

A qualitative positive-or-negative result is sufficient for most uses of rapid infection assays. Even when the test provides quantification (e.g., C_t_ value in PCR), thresholds are usually applied to determine infection status, and patients are usually given the same treatment regardless of the C_t_ value, especially in initial diagnosis. Nonetheless, analyte quantification is useful for some applications, and we were concerned that the number of BAM complexes in the patient samples would not remain linear through all ranges when compared to the PCR C_t_ values. To test this, an additional 10 μL of 6 ng/mL N-protein was spiked in the patient saliva samples. Their fitment into the expected linear regression was then analyzed.

Given the difficulty of counting more than ∼200 BAM complexes due to overlapping tracks and non-linearities due to aggregation and saturation, quantification requires diluting the specimen until it is in a linear range. We thus acquired n=8 PCR-positive specimens, with C_t_ values ranging between 18 and 36 (2.51×10^4^ –0.7 cp/uL according to prior calibrations) and performed a BAM assay on each specimen. If the test was positive, the specimen was serially diluted by 10 or 100x and tested again until diluted to a point where the BAM counts were in the linear range, between the LOD (61 counts) and ∼200 counts. This required up to 10^6^-fold dilution for the concentrated specimens. After dilution to the linear range, the samples were tested in triplicate and their results were compared to the RNA concentrations calculated based on the sample’s C_t_ value. After these analyses, the specimens were further diluted by a factor of 100x (up to 10^8^-fold overall) to see what the background was with almost no analyte. The concentration for each trial was quantified by subtracting the average background signal (20 BAM complexes) from the BAM counts in the linear region and then dividing by the dilution factor. This method is conceptually similar to quantitative PCR, where DNA concentration is determined based on when the fluorescence signal crosses and remains above a threshold. In our case, however, we serially dilute the specimen until the BAM signal falls below the threshold and stays there.

## Supporting information

Data Sheet 1,Data Sheet 2,Data Sheet 3

M1 positive and negative patient BAM assays 60x speed

M2 cell phone movie of 10 uL x 0 and 1 fgmL N-protein –10x speed

## Data Availability

All data will be available in the published version

## Acknowledgements

This research was supported by “Buoyant and Magnetic (BAM) Assays for On Site Sensitive Rapid Diagnostics,” COVID Testing Research Seed Grant Program from Clemson University and “Rising to the Separations and Diagnostic Challenge with Buoyancy and Magnetism (BAM),” Agilent Technologies Inc., Applications and Core Technology – University Relations (ACT-UR).

## Author contributions

The authors had the following CRediT contributor roles: CW: Investigation, methodology, software, visualization, writing original draft and review & editing, ES: investigation, validation, writing reviewing & editing, CL: funding acquisition, writing editing & reviewing; DD: methodology, investigation, funding acquisition, validation, writing editing & reviewing; JNA: conceptualization, formal analysis, methodology, project administration, software, supervision, visualization, funding acquisition, writing original draft and editing & reviewing.

## Supplementary Information

Supplementary Figures and Tables.

Supplementary Movies M1 and M2

