## Supplementary Figure 1-11, Supplementary Table 1-4 for "Rising to the Ultrasensitive Rapid Diagnostic Challenge with Buoyant-Analyte-Magnetic (BAM) Assays"

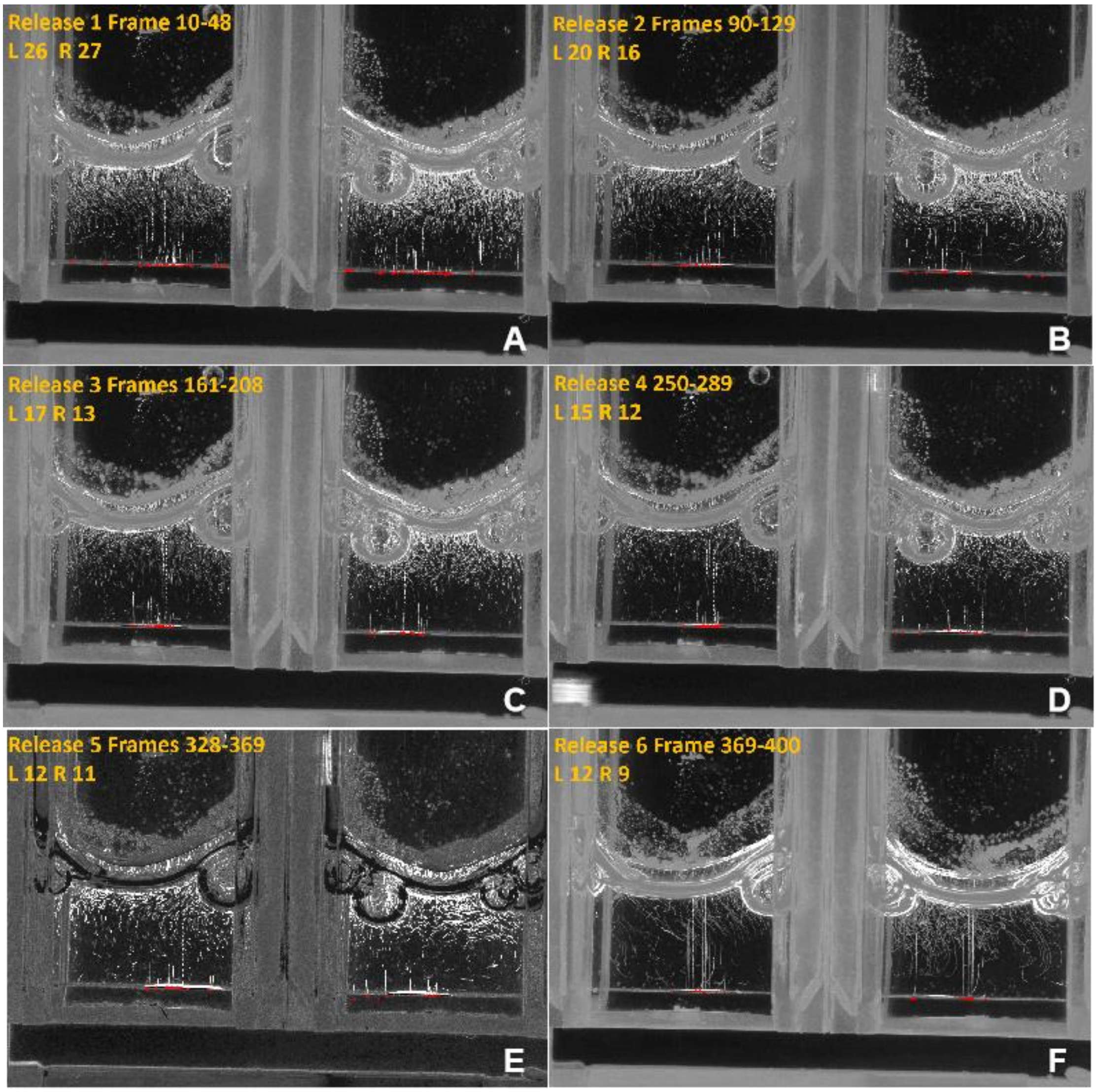

**Figure S1**: The figure A to F showed the gradual reduction of background BAM complexes during six repeated pull-downs by magnet, decreasing from 27 to 11. The sample used is negative sample 7 from Table S4.

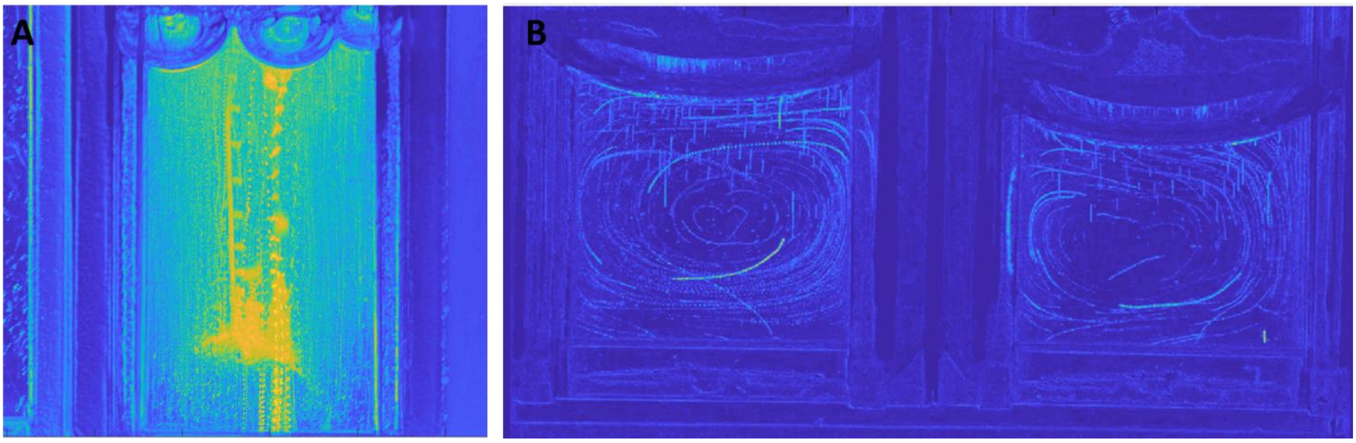

**Figure S2:** Comparison between background reading for negative tests **A)** before additional 2.7 µm magnetic microsphere functionalization **B)** After NSB-reducing procedure. Two adjacent cuvettes with replicate negative samples are imaged at the same time. Tracks that start at the bottom of the cuvette are the nonspecific bound BAM complexes. The arc-shaped trajectory in the middle of the solution are the tracks left by circulating background microbubbles and are distinguished from BAM complexes in not starting at the bottom when the magnet is released. Both cuvettes have 5 nonspecifically bound BAM complexes.

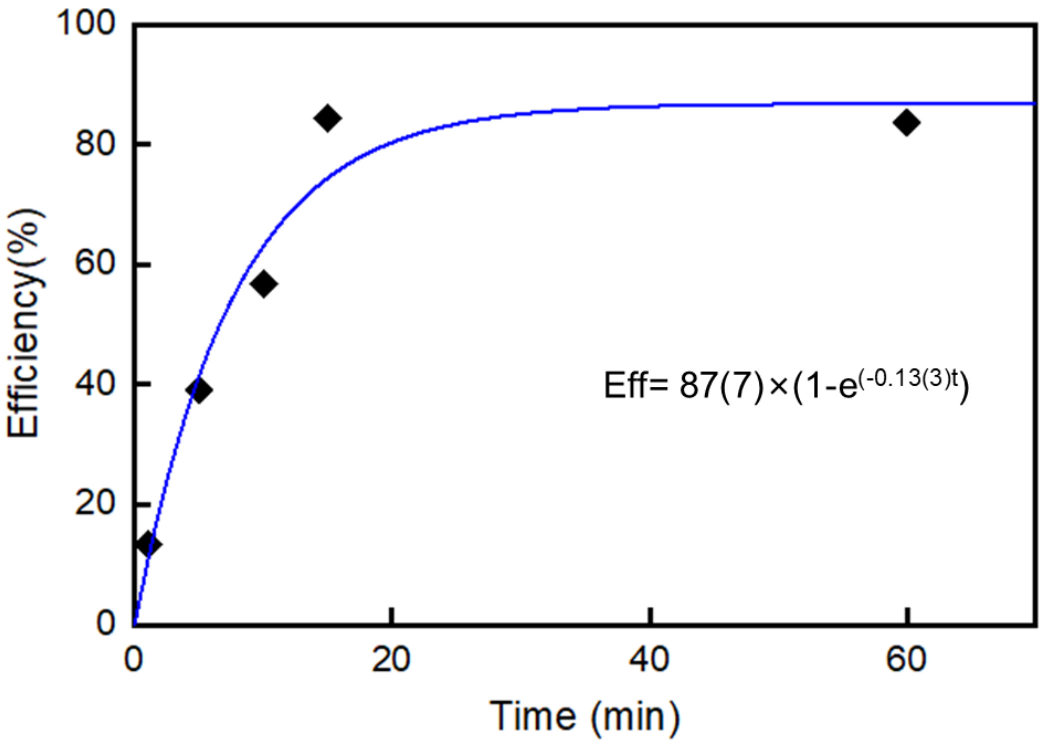

**Figure S3:** BAM capture efficiency vs. incubation period, fit to an exponential regression with 0.13 $min^{-1}$ rate constant (8-minute time constant).

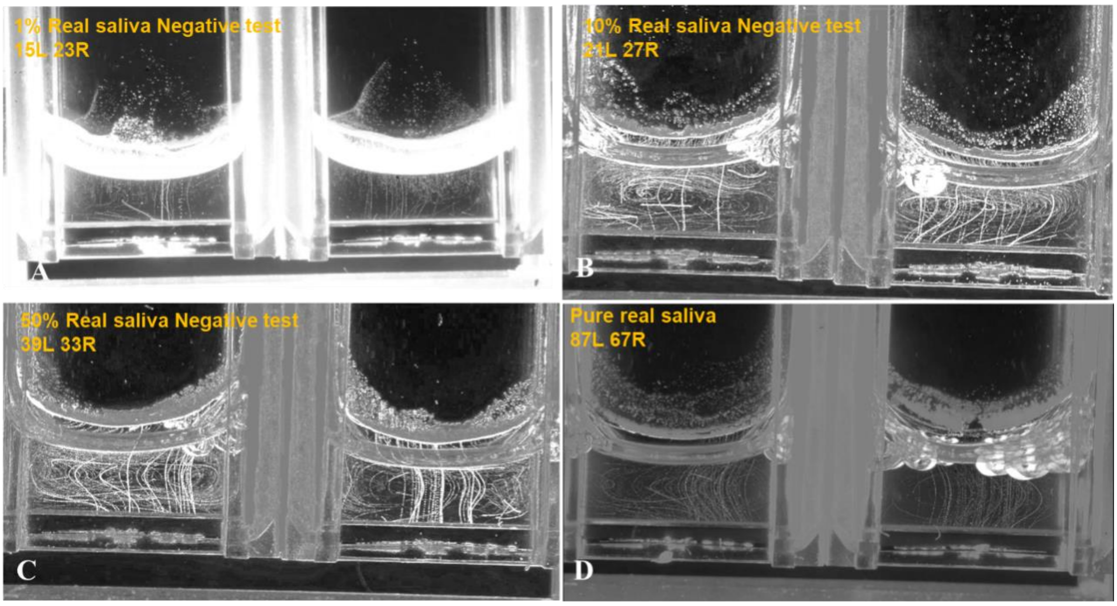

**Figure S4**: BAM tests for varying saliva concentrations. Two adjacent cuvettes with replicate experiments are imaged simultaneously at each concentration. A) 1% saliva negative test: 15 tracks on left, 23 tracks on right. B) 10% saliva negative test: 21 tracks on left, 27 tracks on right. C) 50% saliva negative test: 39 tracks on left, 33 tracks on right. D) Pure saliva negative test: 87 tracks on left, 67 tracks on right

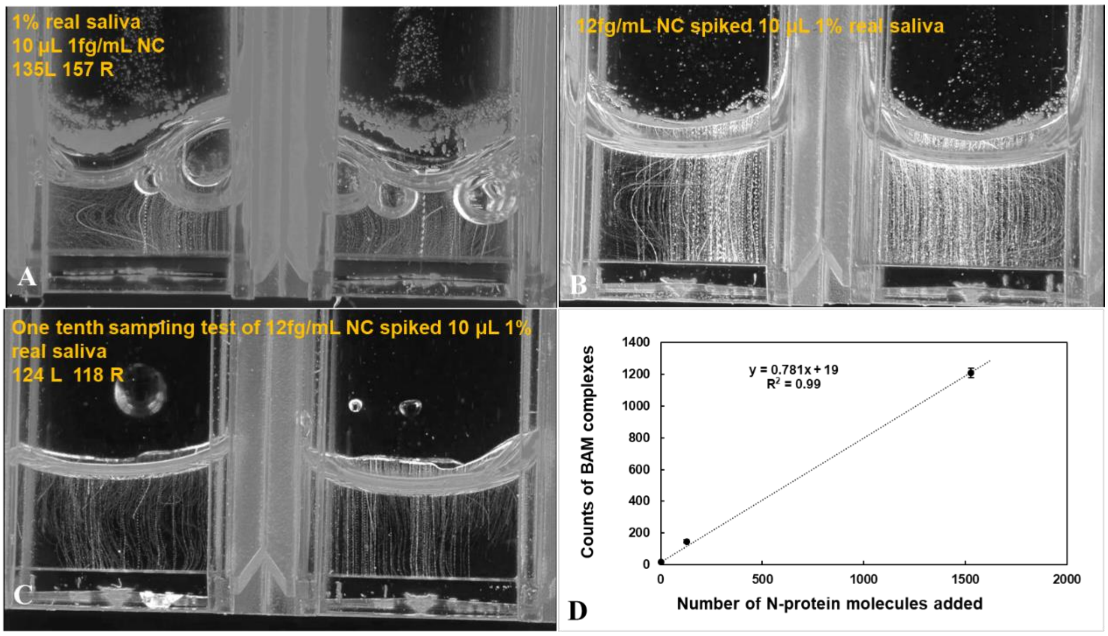

**Figure S5:** BAM tests for N-protein spiked into saliva sample. Two adjacent cuvettes with replicate experiments are imaged simultaneously at each concentration. A) BAM test with 1 fg/mL N-protein (127 molecules) in 1% saliva: 135 BAM tracks on left, 157 on right. B) BAM test with 12 fg/mL N-protein in 1% saliva **C)** One tenth sampling test of B) 124 tracks on left, 118 on right. D) Calibration curve for N-protein BAM test in 1% saliva with 78% capture rate.

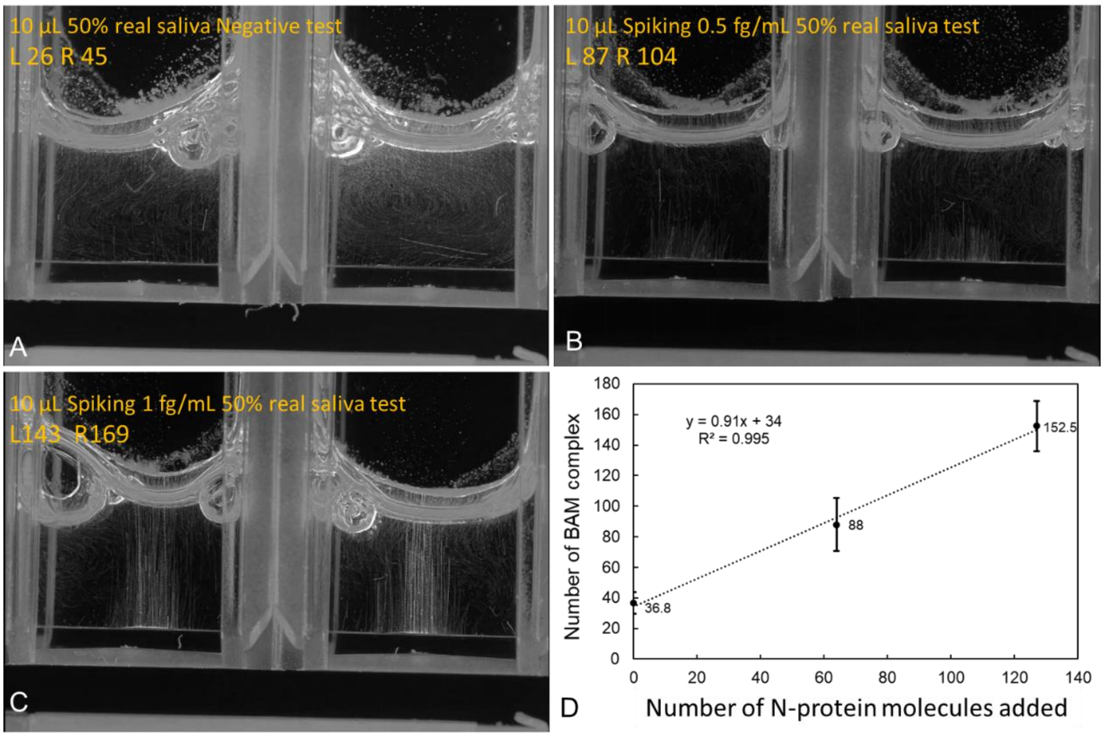

**Figure S6:** BAM tests for 50% saliva concentrations and N-protein spiked saliva sample. Two adjacent cuvettes with replicate experiments are imaged simultaneously at each concentration. A) 50% covid-negative saliva test: 26 tracks on left, 45 on right. B) BAM test with 0.5 fg/mL N-protein (64 molecules) in 50% saliva: 87 tracks on left, 104 on right. C) BAM test with 1 fg/mL N-protein in 50% saliva: 143 tracks on the left 169 tracks on the right. D) Calibration curve for N-protein BAM test in 50% saliva with 91% capture efficiency.

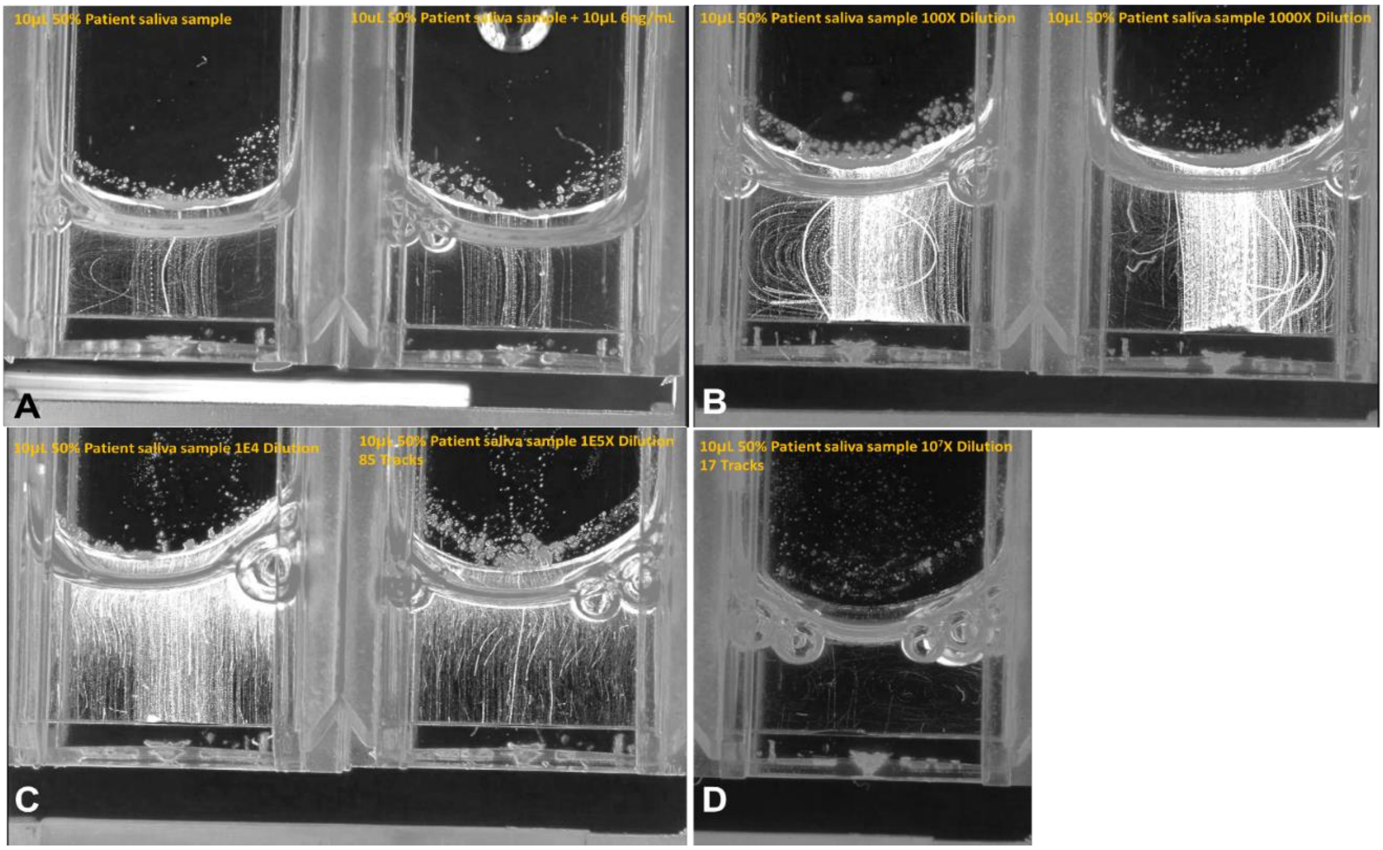

**Figure S7:** Signal changes with analyte spiking of saturated samples and sequential dilution of patient saliva. A) Two adjacent cuvettes imaged simultaneously, the left cuvette had 10 µL 50% patient saliva and the right cuvette had 10 µL 50% patient saliva spiked with 6 ng/mL N-protein. B) The left cuvette contains a 10 µL solution of 50% patient saliva diluted 100-fold, and the right cuvette contains a 10 µL solution diluted 1000-fold. C) The left cuvette contains a 10 µL solution of 50% patient saliva diluted 10,000-fold, and the right cuvette contains a 10 µL solution diluted 100,000-fold, which showed 85 tracks. D) The cuvette contains a 10 µL solution of 50% patient saliva diluted $10^7$-fold.

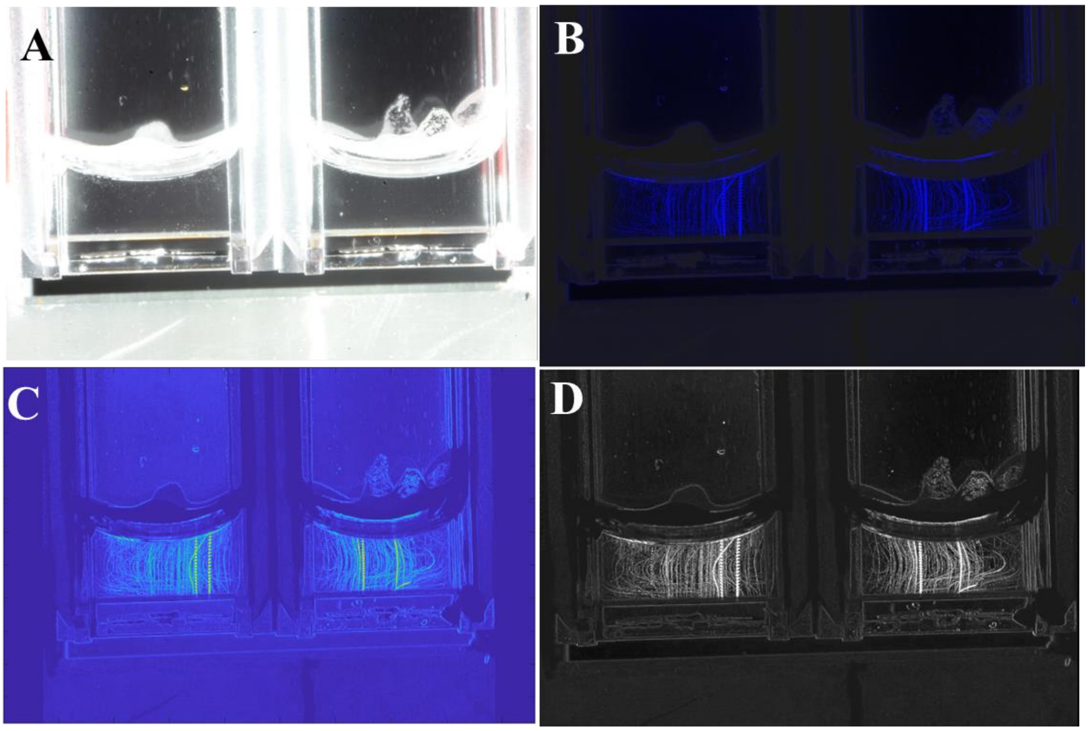

**Figure S8:** Track analysis examples for 127 N-protein spiked simulated saliva, replicated tests in two adjacent cuvettes imaged simultaneously presented in different ways. 127 tracks were observed in the left cuvette and 122 tracks in the right cuvette. (A) Background image. (B) Track analysis colored as blue. (C) Track analysis colored as blue and yellow complementary color. (D) Track analysis grayscale map.

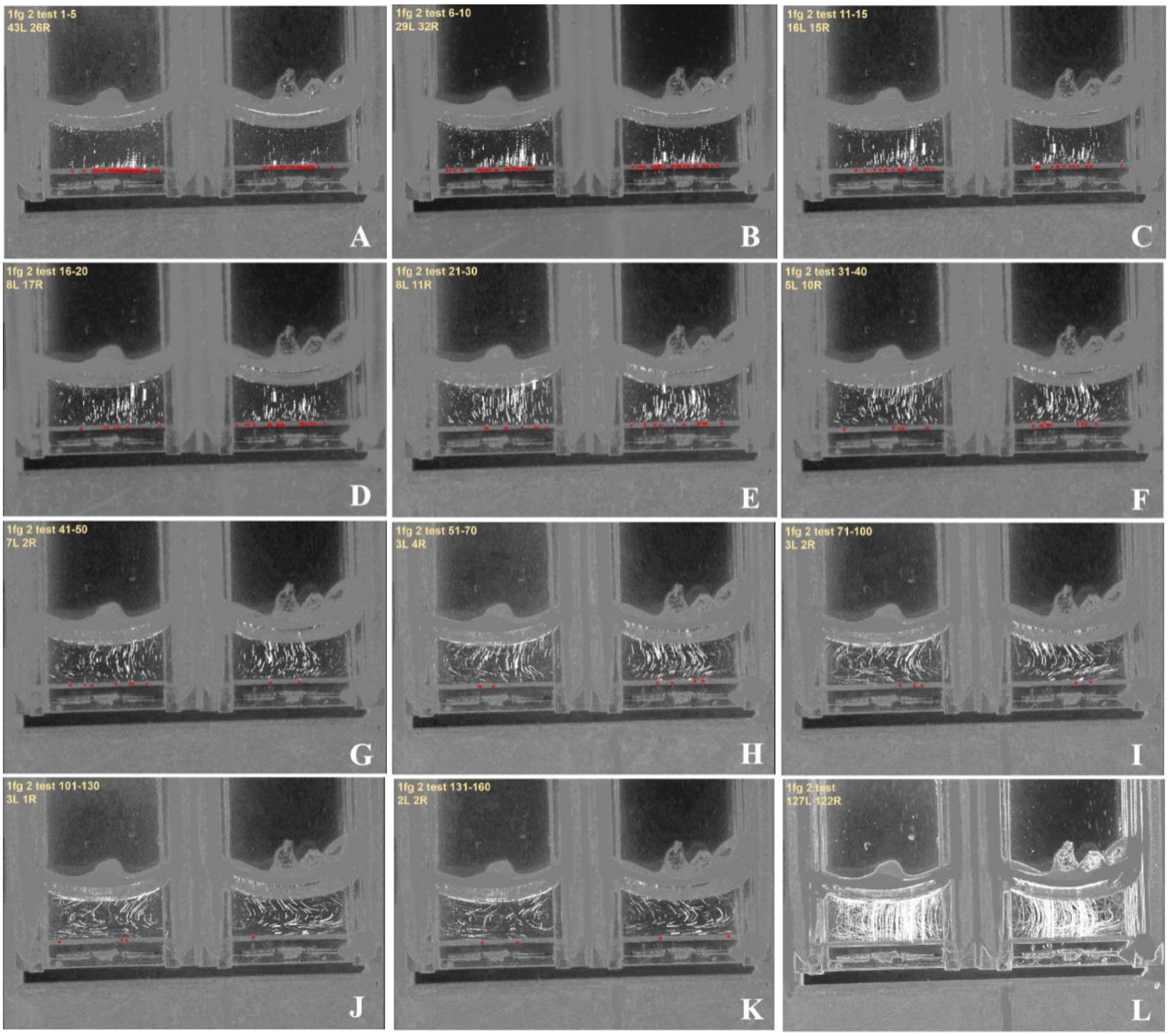

**Figure S9:** Short-period track analysis for BAM test for 10 µL 1 fg/mL N-protein (about 127 molecules). Two adjacent cuvettes with replicate experiments are imaged together. Red stars label new BAM complexes observed. The top left of each subfigure gives the frame numbers in the period and number of BAM complexes. Table S3 shows this data in tabular form.

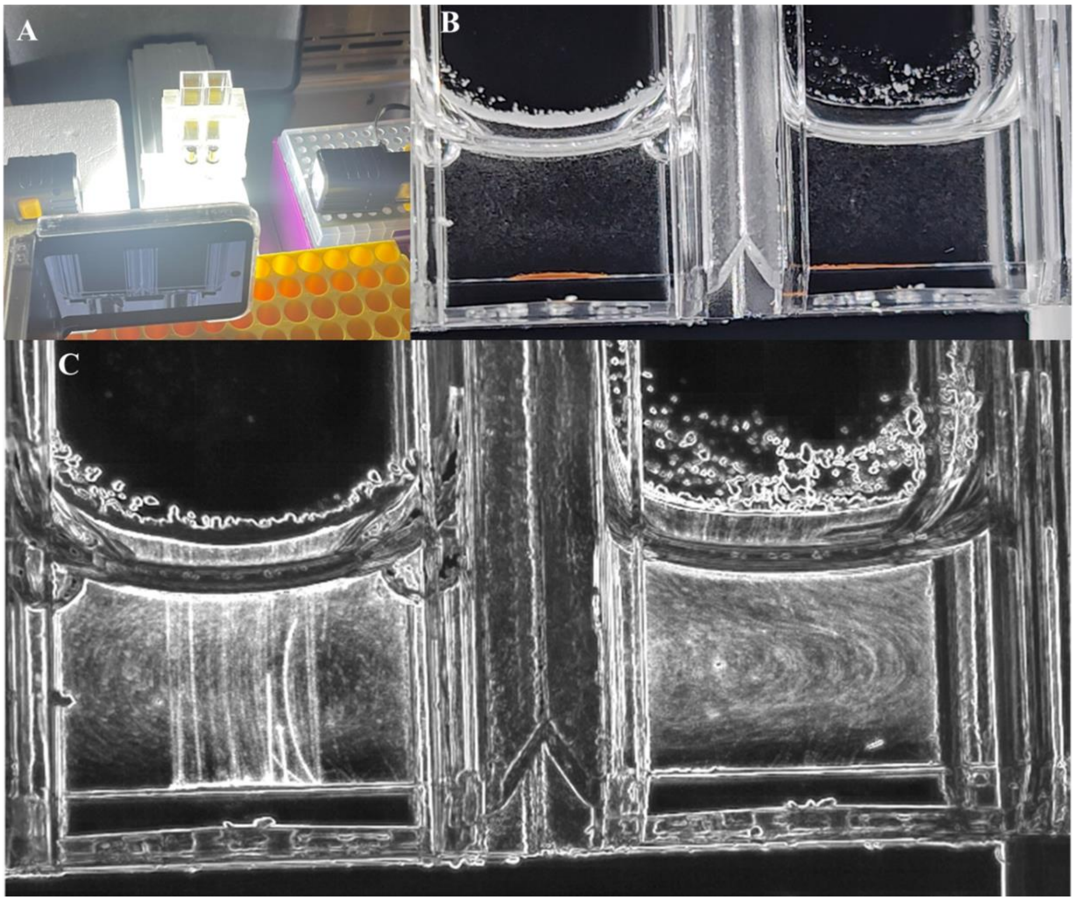

**Figure S10:** BAM test recorded with a cell phone (See Supplementary M2). **A)** The setup contains magnets, 3D printed cuvette holder and magnet holder, two cuvettes, two LED lights, and a Samsung Z Flip phone set to lock focus, 2.4 × zoom, disable auto beautify and filters, and enable HEVC mode. **B)** Real-time image recorded by the phone camera. **C)** Analysis results of the phone video. BAM test with 10 μL 1 × PBS on the right. BAM test with 10 μL 1 fg/mL N-protein on the left.

```
close all;
clear;
clc;
cd('C:\Users\All Users\Example'); %The path to the folder where the video is stored
V1=VideoReader('Example.mp4'); %File name of the video
numFrames = get(V1,'NumberOfFrames'); % The total frame of the video
I=read(V1,1); % Export the first frame of the video
figure (1);
imshow(I);
Io01f1=read(V1,1); % Read the matrix of the first frame
Io01min(:,:)=Io01f1(:,:,1); % Make the first picture as the background and record
% the pixel information in this picture as the maximum and minimum values
Io01max(:,:)=Io01f1(:,:,1); % Make the first picture as the background and record
% the pixel information in this picture as the maximum and minimum values

for i=3:50 % The range of the loop was set to the interval of frame numbers being analyzed

    Io01=read(V1,i); % Take frame into matrix
    display(i); % Show the number of frame in Command Window
    Io01min(:,:)=min(Io01min,Io01(:,:,1));
    Io01max(:,:)=max(Io01max,Io01(:,:,1)); %The matrix obtained form each frame is compared
    % with the background matrix, and the larger value in the matrix is recorded in the Io01max matrix same as min.
end

I1colorb2=0.5*read(V1,3); % Adjust the brightness of the background image
I1colorb2(:,:,3)=I1colorb2(:,:,3) + 1*(Io01max-Io01min); %; Subtracting the matrix of the maximum value from
% the matrix of the minimum value obtains the brightness changes on all pixels,
% and the movement of the BAM complex on a black background could cause such brightness changes.
% So by coloring such brightness changes, the BAM complex movement trajectory displayed.
figure (2);
imshow(I1colorb2); % Plot the track analyses result

figure(3);
imagesc(Io01max-Io01min); % Plot the track analyses result

figure(4);
I1colorb22(:,:)=I1colorb2(:,:,3); % Plot  the track analyses result
imshow(I1colorb22);
colormap(gray);
```

**Figure S11:** MATLAB code for highlighting BAM particle tracks.

| Time / minute | Trial 1 | Trial 2 | Trial 3 | Average number | SD | Background | Effective number | Added N-protein | Efficency |
|---|---|---|---|---|---|---|---|---|---|
| 1 | 49 | 40 | 30 | 39.7 | 9.5 | 22.7 | 17 | 120 | 0.14 |
| 5 | 80 | 78 | 59 | 72.3 | 11.6 | 22.7 | 49.6 | 120 | 0.41 |
| 10 | 102 | 97 | 85 | 94.7 | 8.7 | 22.7 | 72 | 120 | 0.6 |
| 15 | 107 | 153 | 129 | 129.7 | 23 | 22.7 | 107 | 120 | 0.89 |
| 60 | 137 | 125 | 110 | 124 | 13.5 | 17.7 | 106.3 | 120 | 0.89 |

**Table S1:** BAM formation incubation time response.

| Sample | Dilution factor | Average number after dilution | Standard Deviation | Near negative dilution factor | Near negative dilution result | Average for near negative dilution result | Excess BAM counts | Excess BAM counts/μL original saliva | PCR Ct Value | RNA copies/μL | BAM/RNA |
|---|---|---|---|---|---|---|---|---|---|---|---|
| 1 | 2.00E-06 | 90.8 | 23 | 1.00E-08 | 33 | 20.88 | 69.9 | 6992000 | 20.44 | 5214.1 | 1341 |
| 2 | 2.00E-04 | 96.5 | 30 | 1.00E-06 | 15 | | 75.6 | 86420 | 23.60 | 652.6 | 116 |
| 3 | 2.00E-06 | 86.3 | 48 | 1.00E-08 | 10 | | 65.4 | 4282000 | 18.06 | 24859.6 | 263 |
| 4 | 1.00E-06 | 96.3 | 23 | 1.00E-08 | 24 | | 74.4 | 15025000 | 18.38 | 20141.9 | 749 |
| 5 | 1.00E-05 | 70.3 | 11 | 1.00E-07 | 17 | | 49.5 | 989167 | 20.67 | 4467.5 | 221 |
| 6 | 2.00E-01 | 87.0 | 4 | 2.00E-03 | 21 | | 66.1 | 62 | 34.78 | 0.4 | 154 |
| 7 | 1.00E+00 | 234.3 | 40 | 1.00E-02 | 24 | | 213.4 | 40 | 34.02 | 0.7 | 62 |
| 8 | 1.00E-02 | 143.3 | 10 | 1.00E-04 | 23 | | 122.4 | 2449 | 29.18 | 147.7 | 148 |

**Table S2:** Patient diluted saliva BAM test data sheet.

| Figures | A | B | C | D | E | F | G | H | I | J | K | Total |
|---|---|---|---|---|---|---|---|---|---|---|---|---|
| Frames | 1-5 | 6-10 | 11-15 | 16-20 | 21-30 | 31-40 | 41-50 | 51-70 | 71-100 | 101-130 | 131-160 | 1-160 |
| Number of tracks on the left sample | 42 | 29 | 16 | 8 | 8 | 5 | 7 | 3 | 3 | 3 | 2 | 127 |
| Number of tracks on the right sample | 26 | 32 | 15 | 17 | 11 | 10 | 2 | 4 | 2 | 1 | 2 | 122 |

**Table S3:** Counting results of track analysis for BAM test with 10 μL 1 fg/mL N-protein as target. 2-second interval between each frame.

| Reagent | Source | Cat./SKU | Amount | | Unit | | Unit price | Price |
|---|---|---|---|---|---|---|---|---|
| Streptavidin Microbubbles | Akadeum Life Science | 11110-000 | 27 | mL | 2 | pack | $550.00 | $1,100.00 |
| Magnetic bead | Agilent | PL6727-1001 | 3.24 | mL | 2 | pack | $283.00 | $566.00 |
| Ab-R004 | Sino biolocigal | 40143-R004 | 0.0351 | ug | 1 | pack | $400.00 | $400.00 |
| Ab-R040 | Sino biolocigal | 40143-R040 | 0.0351 | ug | 1 | pack | $400.00 | $400.00 |
| Biotinlation-kit | Thermo Scientific | 90407 | 1 | kit | 1 | pack | $379.00 | $379.00 |
| Cuvette | MilliporeSigma | BR759115 | 3600 | each | 36 | pack | $18.70 | $673.20 |
| Centrifuge tube | Thermo Scientific | 90401 | 3600 | each | 15 | pack | $27.13 | $406.95 |
| Salivette | Sarstedt | 51.1534 | 3600 | each | 36 | pack | $60.50 | $2,178.00 |
| misc including PBS, Triton X | | | | | | | | $100.00 |
| | | | | | | | Total | $6,203.15 |
| | | | | | | Price per test | | $1.72 |

**Table S4:** Estimate cost for disposable materials for 3600 tests.
